# A robust phenomenological approach to investigate COVID-19 data for France

**DOI:** 10.1101/2021.02.10.21251500

**Authors:** Q. Griette, J. Demongeot, P. Magal

## Abstract

We provide a new method to analyze the COVID-19 cumulative reported cases data based on a two-step process: first we regularize the data by using a phenomenological model which takes into account the endemic or epidemic nature of the time period, then we use a mathematical model which reproduces the epidemic exactly. This allows us to derive new information on the epidemic parameters and to compute the effective basic reproductive ratio on a daily basis. Our method has the advantage of identifying robust trends in the number of new infectious cases and produces an extremely smooth reconstruction of the epidemic. The number of parameters required by the method is parsimonious: for the French epidemic between February 2020 and January 2021 we use only 11 parameters in total.

## 1 Introduction

Modeling endemic and epidemic phases of the infectious diseases such as smallpox which by the 16th century had become a predominant cause of mortality in Europe until the vaccination by E. Jenner in 1796, and present Covid-19 pandemic outbreak has always been a means of describing and predicting disease. D. Bernoulli proposed in 1760 a differential model [1] taking into account the virulence of the infectious agent and the mortality of the host, which showed a logistic formula [1, p. 13] of the same type as the logistic equation by Verhulst [2]. The succession of an epidemic phase and followed by an endemic phase has been introduced by Bernoulli and for example appears clearly in the Figures 9 and 10 in [3].

The aim in this article is to propose a new approach to compare epidemic models with data from reported cumulative cases. Here we propose a phenomenological model to fit the observed data of cumulative infectious cases of COVID-19 that describe the successive epidemic phases and endemic intermediate phases. This type of problem dates back to 70^th^ with the work of London and York [4]. More recently, Chowel et al. [5] have proposed a specific function to model the temporal transmission speeds *τ* (*t*). In the context of COVID-19, a two-phase model has been proposed by Liu et al. [6] to describe the South Korean data with an epidemic phase followed by an endemic phase.

In this article, we use a phenomenological model to fit the data (see Figure 1). The phenomenological model is used in the modelling process between the data and the epidemic models. The difficulty here is to propose a simple phenomenological model (with limited number of parameters) that would give a meaningful result for the time dependent transmission rates *τ* (*t*). Many models could potentially be used as phenomenological to represent the data (ex. cubic spline and others). The major difficulty here is to provide a model that gives a good description of the tendency for the data. It has been observed in our previous work that it is difficult to choose between the possible phenomenological models (see Figures 12-14 in [8]). The phenomenological model can also be viewed as a regularization of data that should not fluctuate too much in order to keep the essential information. An advantage in our phenomenological model is the limited number of parameters (5 parameters during each epidemic phase and 2 parameters during each endemic phase). The last advantage of our approach is that once the phenomenological model has been chosen, we can compute some explicit formula for the transmission rate and derive some estimations for the other parameters.

**Figure 1:**
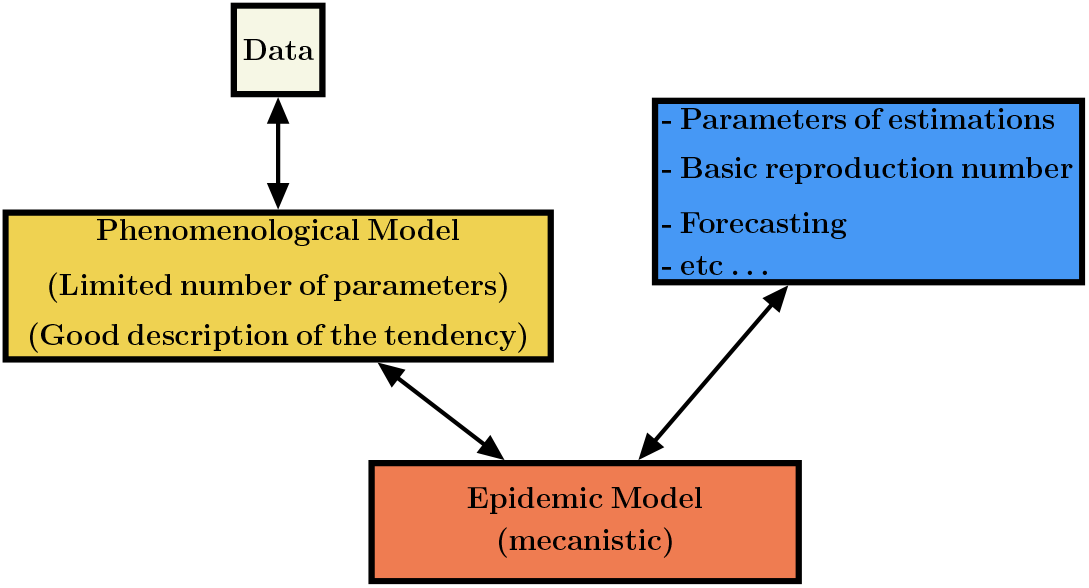
We can apply statistical methods to estimate the parameters of the proposed phenomenological model and derive their average values with some confidence intervals. The phenomenological model is used at the first step of the modelling process, providing regularized data to the epidemic model and allowing the identification of its parameters.

## 2 Material and methods

### 2.1 Phenomenological model

In this article, the phenomenological model is compared with the cumulative reported cases data taken from WHO [7]. The phenomenological model deals with data series of new infectious cases decomposed into two types of successive phases, 1) endemic phases, followed by 2) epidemic ones.

#### Endemic phase

During the endemic phase the dynamics of new cases appears to fluctuate around an average value independently of the number of cases. Therefore the on average cumulative number of cases is given by

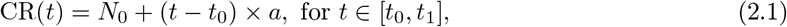

where *t*_0_ denotes the beginning of the endemic phase. *a* is the average value of CR(*t*_0_) and *N*_0_ the average value of the daily number of new cases.

In other words, we assume that the average daily number of new cases is constant. Therefore the daily number of new cases is given by

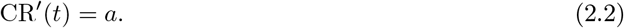

#### Epidemic phase

In the epidemic phase, the new cases are contributing to produce second cases. Therefore the daily number of new cases is no longer constant but varies with time as follows

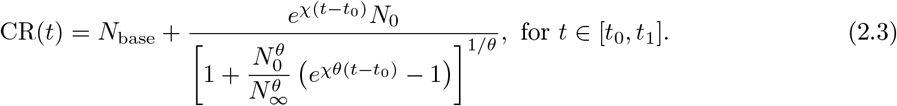

In other words, the daily number of new cases follows the Bernoulli-Verhulst [1, 2] equation. Namely, by setting *N* (*t*) = CR(*t*) *− N*_base_ we obtain

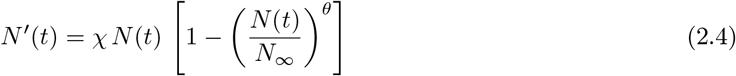

completed with the initial value

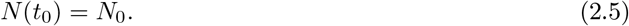

In the model *N*_base_ + *N*_0_ corresponds to the value CR(*t*_0_) of the cumulative number of cases at time *t* = *t*_0_. The parameter *N*_*∞*_ + *N*_base_ is the maximal increase of the cumulative reported cases after the time *t* = *t*_0_. *χ >* 0 is a Malthusian growth parameter, and *θ* regulates the speed at which the CR(*t*) increases to *N*_*∞*_ + *N*_base_.

#### Regularized model

Because the formula for *τ* (*t*) involves derivatives of the phenomenological model regularizing CR(*t*) (see equation (2.12)), we need to connect the phenomenological models of the different phases as smoothly as possible. We let 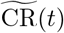 be the model obtained by placing the phenomenological models for the different phases side by side. Outside of the time window where phenomenological models are used, we consider that the function 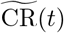 is constant. We define the regularized model by using the convolution formula:

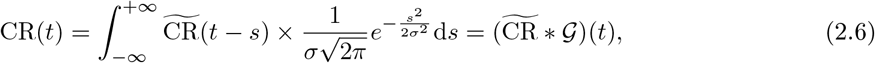

where 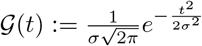 is the Gaussian function of variance *σ*^2^. The parameter *σ* controls the trade-off between smoothness and precision: increasing *σ* reduces the variations in CR(*t*) and reducing *σ* reduces the distance between CR(*t*) and 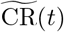. In any case the resulting function CR(*t*) is very smooth (as well as its derivatives) and close to the original model 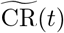 when *σ* is not too large. In numerical applications, we take *σ* = 2 days.

#### Procedure to fit the phenomenological model to the data

In order to fit the model to the data, we used the regularized model (2.6) where the periods of the different phases are fixed as in Table 1. We use a standard curve-fitting algorithm to find the parameters of the regularized model. In numerical applications we used the Levenberg–Marquardt nonlinear least squares algorithm provided by the MATLAB_©_ function fit. Our 95% confidence intervals are the ones provided as an output of this algorithm. The best fit parameters and the corresponding confidence intervals are provided in Table 1.

**Table 1:** Fitted parameters and computed parameters for the whole epidemic.

| Period | Parameters value | Method | 95% Confidence interval |
| --- | --- | --- | --- |
| Period 1: Endemic phase<br>Jan 03 - Feb 27 | $N_0 = -4.368$<br>$a = 1.099 \times 10^{-1}$ | computed<br>fitted | $a \in [-8.582 \times 10^1, 8.604 \times 10^1]$ |
| Period 2: Epidemic phase<br>Feb 27 - May 17 | $N_{base} = 0$<br>$N_0 = 1.675$<br>$N_\infty = 1.445 \times 10^5$<br>$\chi = 1.263$<br>$\theta = 6.315 \times 10^{-2}$ | fixed<br>fitted<br>fitted<br>fitted<br>fitted | $N_0 \in [-3.807 \times 10^1, 4.142 \times 10^1]$<br>$N_\infty \in [1.367 \times 10^5, 1.523 \times 10^5]$<br>$\chi \in [-1.171 \times 10^1, 1.424 \times 10^1]$<br>$\theta \in [-6.086 \times 10^{-1}, 7.349 \times 10^{-1}]$ |
| Period 3: Endemic phase<br>May 17 - Jul 05 | $N_0 = 1.405 \times 10^5$<br>$a = 3.11 \times 10^2$ | computed<br>computed | |
| Period 4: Epidemic phase<br>Jul 05 - Nov 18 | $N_{base} = 1.403 \times 10^5$<br>$N_0 = 1.517 \times 10^4$<br>$N_\infty = 1.953 \times 10^6$<br>$\chi = 3.671 \times 10^{-2}$<br>$\theta = 7.679$ | fitted<br>fitted<br>fitted<br>fitted<br>fitted | $N_{base} \in [1.367 \times 10^5, 1.439 \times 10^5]$<br>$N_0 \in [1.427 \times 10^4, 1.607 \times 10^4]$<br>$N_\infty \in [1.92 \times 10^6, 1.986 \times 10^6]$<br>$\chi \in [3.62 \times 10^{-2}, 3.722 \times 10^{-2}]$<br>$\theta \in [6.256, 9.102]$ |
| Period 5: Endemic phase<br>Nov 18 - Jan 04 | $N_0 = 4.45 \times 10^{-84}$<br>$a = 1.099 \times 10^{-1}$ | computed<br>fitted | $a \in [1.222 \times 10^4, 1.265 \times 10^4]$ |

### 2.2 SI Epidemic model

The SI epidemic model used in this work is the same as in [8]. It is summarized by the flux diagram in Figure 2.

**Figure 2:**
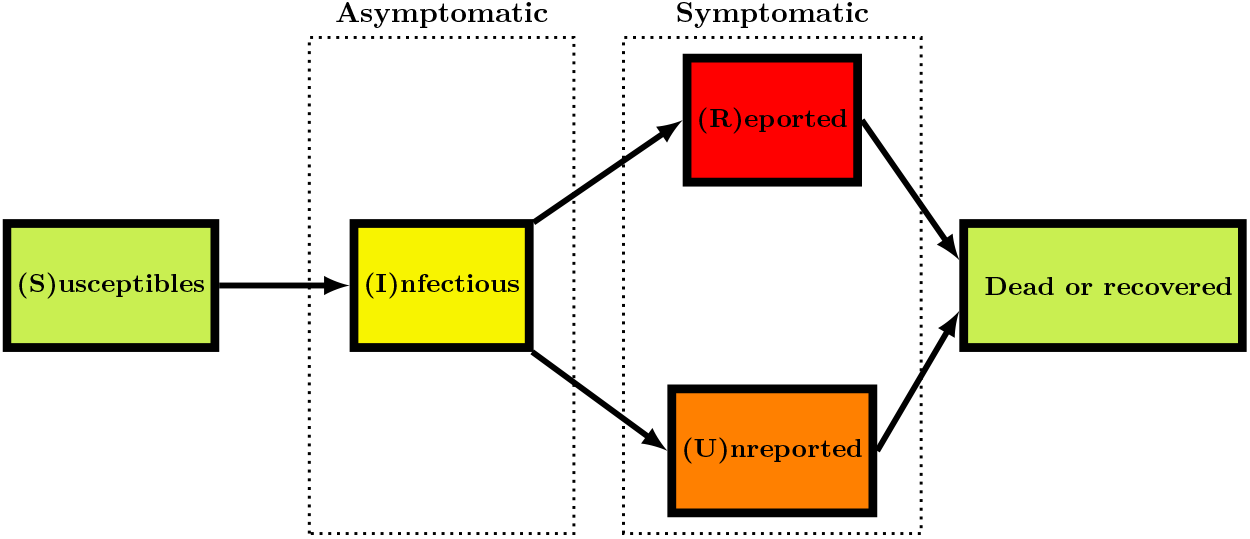
Schematic view showing the different compartments and transition arrows in the epidemic model.

The goal of this article is to understand how to compare the SI model to the reported epidemic data and therefore the model can be used to predict the future evolution of epidemic spread and to test various possible scenarios of social mitigation measures. For *t ≥ t*_0_, the SI model is the following

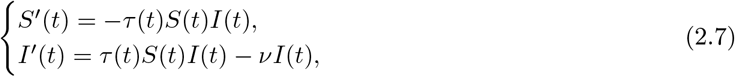

where *S*(*t*) is the number of susceptible and *I*(*t*) the number of infectious at time *t*. This system is supplemented by initial data

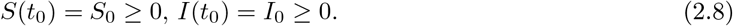

In this model, the rate of transmission *τ* (*t*) combines the number of contacts per unit of time and the probability of transmission. The transmission of the pathogen from the infectious to the susceptible individuals is described by a mass action law *τ* (*t*) *S*(*t*) *I*(*t*) (which is also the flux of new infectious).

The quantity 1*/ν* is the average duration of the infectious period and *νI*(*t*) is the flux of recovering or dying individuals. At the end of the infectious period, we assume that a fraction *f ∈* (0, 1] of the infectious individuals is reported. Let CR(*t*) be the cumulative number of reported cases. We assume that

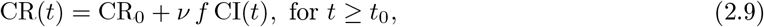

Where

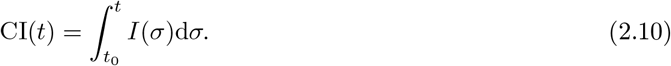

#### Assumption 2.1 (Given parameters) *We assume that*

- *the number of susceptible individuals when we start to use the model S*_0_ = 67 *millions;*
- *the average duration of infectious period* 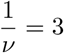 *days;*
- *the fraction of reported individuals f* = 0.9; *are known parameters*.

#### Parameters estimated in the simulations

As described in [8] the number of infectious at time *t*_0_ is

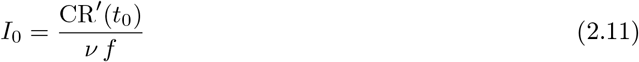

The rate of transmission *τ* (*t*) at time *t* is given by

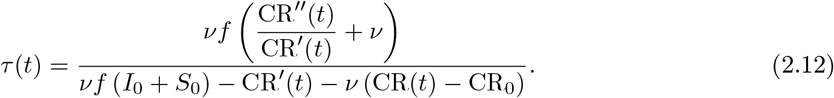

#### Parameters estimated in the endemic phase

The initial number of infectious is given by

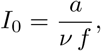

and the transmission rate is given by the explicit formula

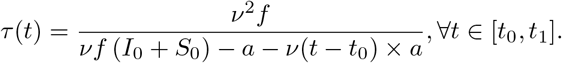

#### Parameters estimated in the epidemic phase

The initial number of infectious is given by

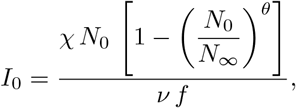

and the transmission rate is given by the explicit formula

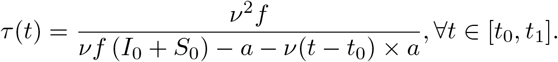

and since

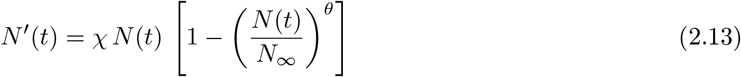

and

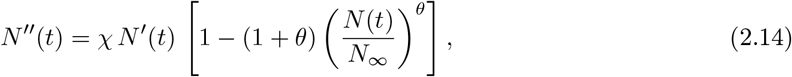

we obtain an explicit formula

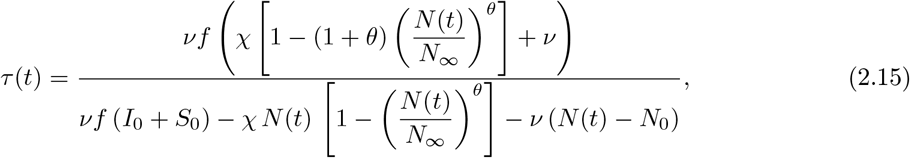

with

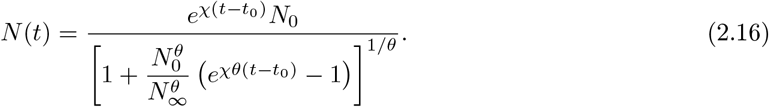

By using the Bernoulli-Verhulst model to represent the data, the daily number of new cases is nothing but the derivative *N* ^*I*^(*t*) (whenever the unit of time is one day). The daily number of new cases reaches its maximum at the turning point *t* = *t*_*p*_, and by using (2.14), we obtain

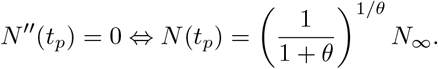

Therefore by using (2.13) the maximum of the daily number of cases equals

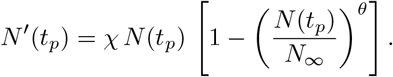

By using the above formula, we obtain a new indicator for the amplitude of the epidemic.

**Theorem 2**.**2** *The maximal daily number of cases in the course of the epidemic phase is given by*

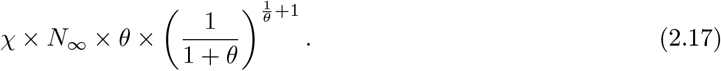

### Parameter bounds

The epidemic model (2.7) with time-dependent transmission rate is consistent only insofar as the transmission rate remains positive. This gives us a criterion to judge if a set of epidemic parameters has a chance of being consistent with the observed data: since we know the parameters *N*_0_, *N*_*∞*_, *χ* and *θ* from the phenomenological model, the formula (2.18) allows us to compute a criterion on *ν* and *f* which decides whether a given parameter values are compatible with the observed data or not. That is to say that, a set of parameter values is compatible if the transmission rate *τ* (*t*) in (2.18) remains positive for all *t ≥t*_0_, and it is not compatible if the sign of *τ* (*t*) in (2.18) changes for some *t ≥t*_0_.

The value of the parameter *ν* is compatible with the model (2.18) if and only if

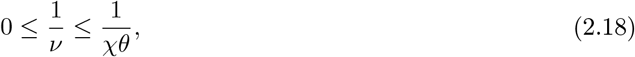

and the value of the parameter *f* is compatible with the model (2.18) if and only if

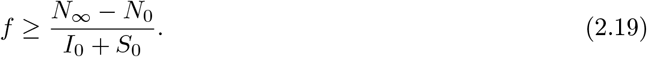

Therefore, we obtain an information on the parameters *ν* and *f*, even though they are not directly identifiable (two different values of *ν* or *f* can produce exactly the number same cumulative reported cases).

### 2.4 Computation of the basic reproduction number

In order to compute the reproduction number in Figure 5 with use the Algorithm 2 in [8] and the day by day values of the phenomenological model.

## 3 Results

### 3.1 Phenomenological model compared to the French data

In Figure 3 we present the best fit of our phenomenological model for the cumulative reported cases data of COVID-19 epidemic in France. The yellow regions correspond to the endemic phases and the blue regions correspond to the epidemic phases. Here we consider the two epidemic waves for France, and the chosen period as well as the parameters values for each period are listed in Table 1. In Table 1 we also give 95% confidence intervals for the parameters values.

**Figure 3:**
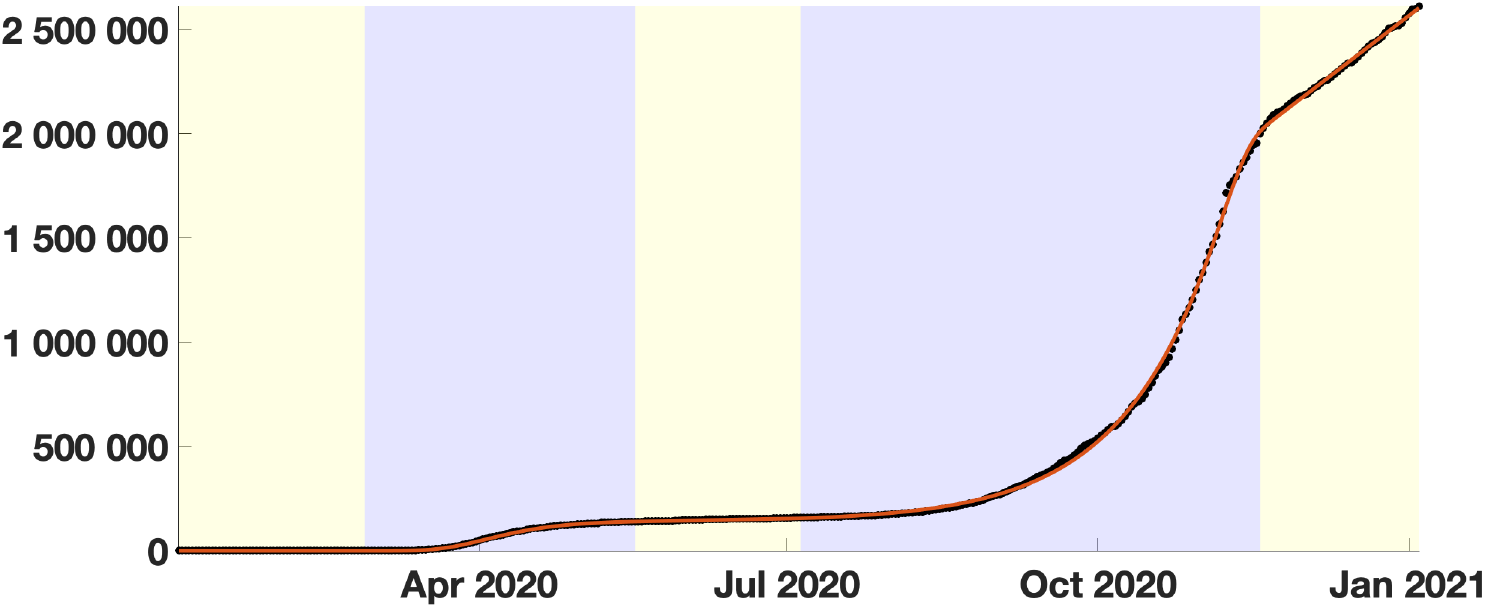
The red curve corresponds to the phenomenological model and the black dots correspond to the cumulative number of reported cases in France.

Figure 4 shows the corresponding daily number of new reported cases data (black dots) and the first derivative of our the phenomenological model (red curve).

**Figure 4:**
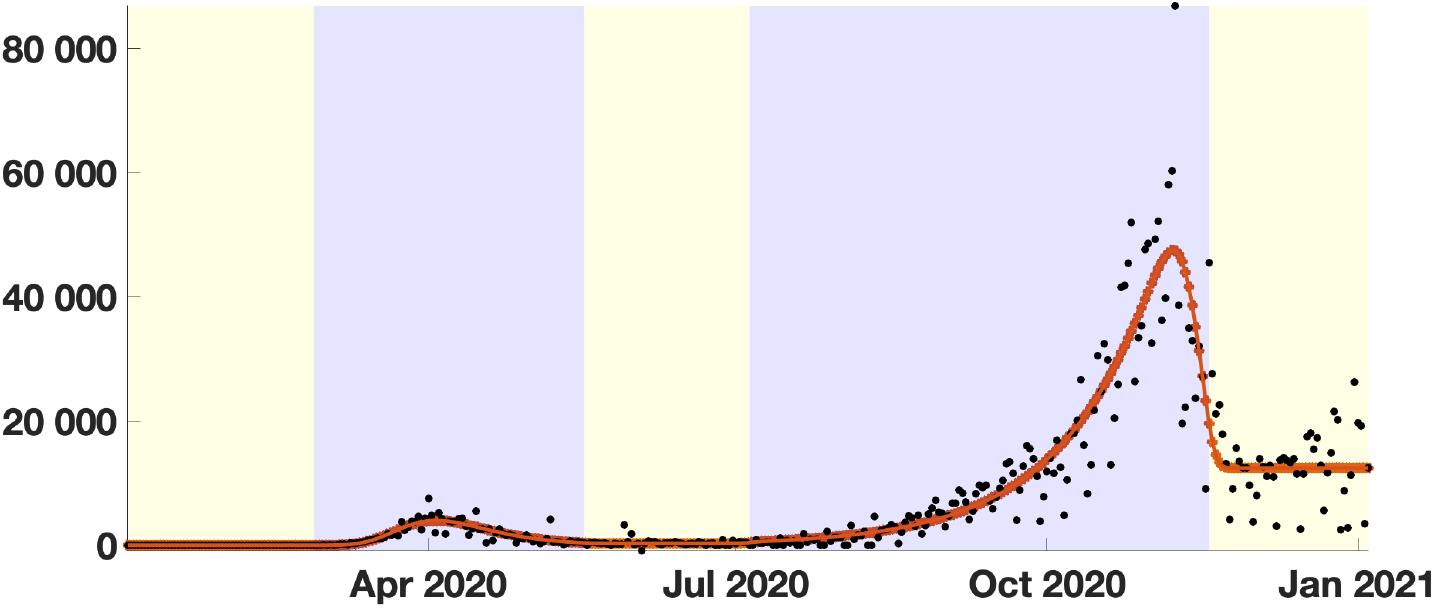
The red curve corresponds to the first derivative of the phenomenological model and the black dots correspond to daily number of new reported cases in France.

**Figure 5:**
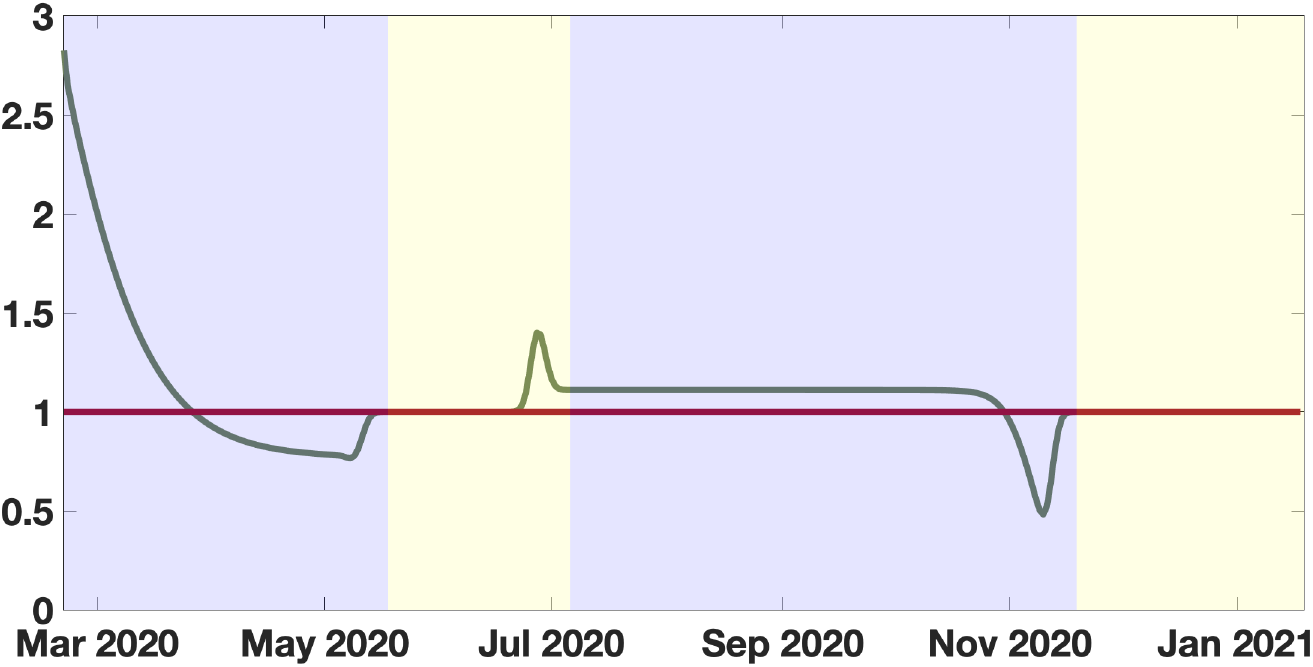
In this figure we plot the time dependent basic reproduction number ℛ_0_(t) := τ (t)S(t)/ν. We fix the average length of the asymptotic infectious period to 3 days.

### 3.2 SI epidemic model compared to the French data

Some parameters of the model are known like *S*_0_ = 67 millions for France (this is questionable). Some parameters of the epidemic model can not be precisely evaluated [8].

#### Result

By using (2.18) we obtain the following conditions for the average duration of infectious period

- 0 < 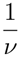 ≤ 1/(*χθ*) = 12.5 days during the first epidemic wave;
- 0 < 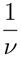 ≤ 1/(*χθ*) = 3.5 days during the second epidemic wave.

We obtain no constraint for the fraction *f* ∈ (0, 1] of reported new cases (between 0 and 1 for France).

Moreover by using the formula (2.17) we deduce that the maximal daily number of cases is

- 4110 during the first epidemic wave;
- 47875 during the second epidemic wave.

Importantly, by combining the phenomenological model from Section 3.1 and the epidemiological model from Section 2.2, we can reconstruct the time-dependent transmission rate given by (2.12) and the corresponding time-dependent basic reproduction number ℛ_0_(*t*) = *τ* (*t*)*S*(*t*)*/ν* (sometimes called “effective basic reproductive ratio”). The obtained basic reproduction number if presented in Figure 5. We observe that ℛ_0_(*t*) is decreasing during each epidemic wave, except at the very end where it becomes increasing. This is not necessarily surprising since the lockdown becomes less strictly respected towards the end. During the endemic phases, the ℛ_0_(*t*) becomes effectively equal to one, except again near the end. The variations observed close to the transition between two phases may be partially due to the smoothing method, which has an impact on the size of the “bumps”. However they remain very limited in number and size.

## 4 Discussion

In our paper we use a phenomenological model to reduce the number of the parameters necessary for summarizing observed data without loss of pertinent information. The process of reduction consists of three stages: qualitative or quantitative detection of the boundaries between the different phases of the dynamics (here endemic and epidemic phases), choice of a reduction model (among different possible approaches: logistic, regression polynomials, splines, autoregressive time series, etc.) and smoothing of the derivatives at the boundary points corresponding to the breaks in the model.

In Figure 3, we have a very good agreement between the data and the phenomenological model, for both the original curve and its derivative. The relative error in Figure 3 is of order 10^−2^, which means that the error is at most of order of 100 000 individuals. In Figure 4 the red curve also gives a good tendency of the black dots corresponding to raw data.

In Figure 5, the phenomenological models are necessary to derive a significant basic reproduction number. Otherwise the resulting *ℛ*_0_(*t*) is not interpretable and even not computable after sometime. Similar results were obtained in Figures 12-14 in [8]. The method to compute *ℛ*_0_ can also be applied directly to the original data. We did not show the result here because the noise in the data is amplified by the method and the results are not usable. This shows that it is important to use the phenomenological model to provide a good regularization of the data.

In Figure 5, the major difficulty is to know how to make the transition from an epidemic phase to an endemic phase and vice versa. This is a non-trivial problem which is solved by our regularization approach (using a convolution with a Gaussian). As we can see in Figure 5, the number of oscillations is very limited between two phases. Without regularization, there is a sharp corner at the transition between two phases which leads to infinite values in *τ* (*t*). The choice of the convolution with a Gaussian kernel for the regularization method is the result of an experimental process. We tried several different regularization methods, including a smooth explicit interpolation function and Hermite polynomials. Eventually, the convolution with a Gaussian kernel gives the best results.

In order to minimize the variations of the curve of *ℛ*_0_(*t*), the choice of the transition dates between two phases is critical. In Figure 5 we choose the transition dates so that the derivatives of the phenomenological model do not oscillate too much. Other choices lead to higher variations or increase the number of oscillations. Finally, the qualitative shape of the curve presented in Figure 5 is very robust to changes in the epidemic parameters, even though the quantitative values of *ℛ*_0_(*t*) are different for other values of the parameters *ν* and *f*.

In Figure 5, we observe that the quantitative value of the *ℛ*_0_(*t*) during the first part of the second epidemic wave (second blue region) is almost constant and equals 1.11. This value is significantly lower than the one observed at the beginning of the first epidemic wave (first blue region). Yet the number of cases produced during the second wave is much higher than the number of cases produced during the first wave.

We observe that the values of the parameters of the phenomenological model are quantitatively different between the first wave and the second wave. Several phenomena can explain this difference. The population was better prepared for the second wave. The huge difference in the number of daily reported cases during the second phase can be partially attributed to the huge increase in the number of tests in France during this period. But this is only a partial explanation for the explosion of cases during the second wave. We also observe that the average duration of infectious period varies between the first epidemic wave (12.5 days) and the second epidemic wave (3.5 days). This may indicate a possible adaptation of the virus SARS CoV-2 circulating in France during the two periods, or the effect of the mitigation measures, with a better respect of the social distancing and compulsory mask wearing.

The huge difference between the initial values of *ℛ*_0_(*t*) in the first and the second waves is an apparent paradox which shows that *ℛ*_0_(*t*) has a limited explanatory value regarding the severity of the epidemic: even if the quantitative value of *ℛ*_0_(*t*) is higher at the start of the first wave, the number of cases produced during an equivalent period in the second wave is much higher. This paradox can be partially resolved by remarking that the *ℛ*_0_(*t*) behaves like an exponential rate and the number of secondary cases produced in the whole population is therefore very sensitive to the number of active cases at time *t*. In other words, *ℛ*_0_(*t*) is blind to the epidemic state of the population and cannot be used as a reliable indicator of the severity of the epidemic. Other indicators have to be found for that purpose; we propose, for instance, the maximal value of the daily number of new cases, which can be forecasted by our method (see equation (2.17)), although other indicators may possibly be imagined.

In Figure 6 we present an exploratory scenario assuming that during the endemic period preceding the second epidemic wave (May 17 - Jul 05) the daily number of cases is divided by 10. The resulting cumulative number of cases obtain is five lower the original one. We summarize this observation into the following statement.

**Figure 6:**
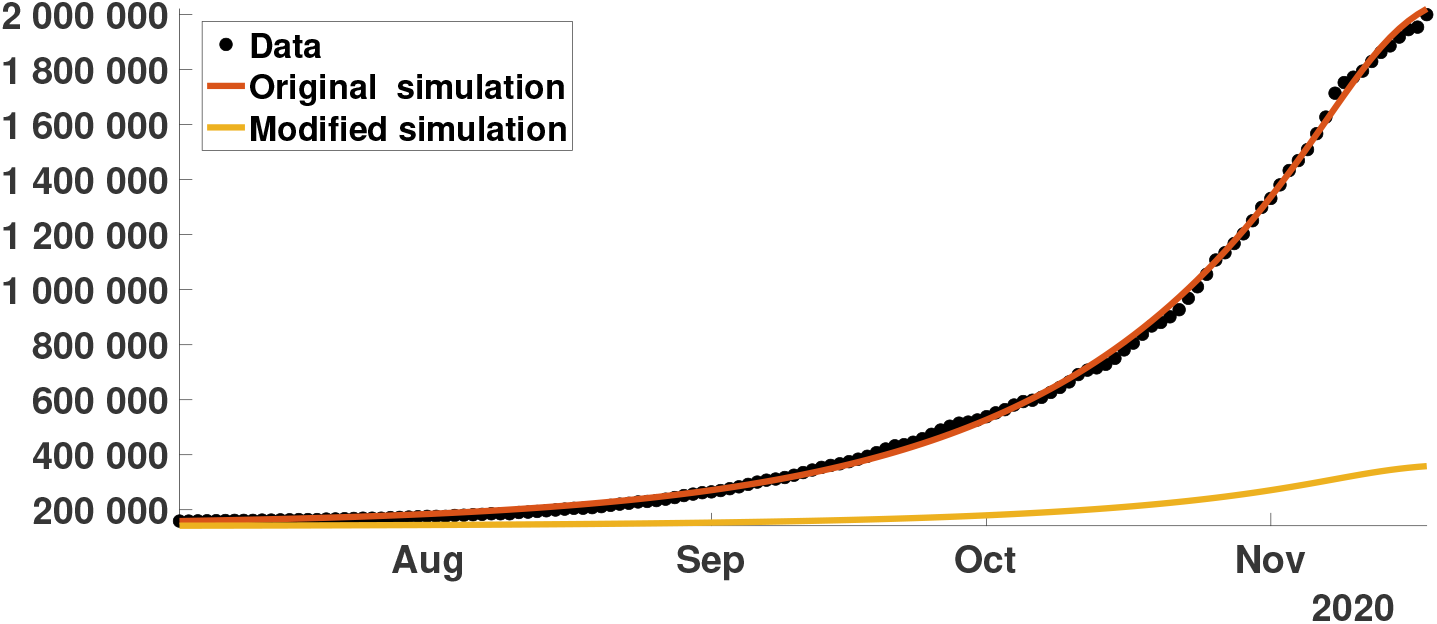
Cumulative number of cases for the second epidemic wave obtained by using the SI model (2.7) with τ (t) given by (2.12), the parameters from Table 1. We start the simulation at time t_0_ = July 05 with the initial value 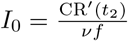 for red curve and with 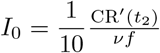for yellow curve. The remaining parameters used are ν = 1/3, f = 0.9, S_0_ = 66841266. We observe the number is five times lower than then the original number of cases.

### Result

- The level of the daily number of cases during an endemic phase preceding an epidemic phase strongly influences the severity of this epidemic wave.
- In other words, maintaining social distancing between epidemic waves is essential.

In Figure 4, there is two order of magnitude in the daily number of cases in between *CR*′(*t*_1_) *≈*1 (with *t*_1_ = Feb 27) at the early beginning of the first epidemic wave and *CR*′(*t*_2_) = 422 (with *t*_2_ = July 05) at the early beginning of the second epidemic wave. That confirms our result. After the second wave, the average daily number of cases *CR*′(*t*) in France is stationary and approximately equal to 12440. Therefore, if the above observation remains true and if a third epidemic wave occurs, the third epidemic wave is expected to be more severe than the first and second epidemic waves.

Our study can be extended in several directions. A statistical study of the parameters obtained by using our phenomenological model with data at the regional scale could be interesting. We could in particular investigate statistically the correlations existing between the parameters changes and the variations with demographic parameters as the median age and the population density, as well as geoclimatic factors as the elevation and temperature, etc. We also plan to extend our method to more realistic epidemic models, like the SEIUR model from [9], which includes the possibility of transmission from asymptomatic unreported patients.

## Data Availability

No data were produced

